# Controlled randomized clinical trial on using Ivermectin with Doxycycline for treating COVID-19 patients in Baghdad, Iraq

**DOI:** 10.1101/2020.10.26.20219345

**Authors:** Hashim A. Hashim, Mohammed F. Maulood, Anwar M. Rasheed, Dhurgham F. Fatak, Khulood K. Kabah, Ahmed S. Abdulamir

**Author notes:** Registered in clinicaltrials.gov: NCT04591600.

## Abstract

**Objectives:** COVID-19 patients suffer from the lack of curative therapy. Hence, there is an urgent need to try repurposed old drugs on COVID-19.

**Methods:** Randomized controlled study on 70 COVID-19 patients (48 mild-moderate, 11 severe, and 11 critical patients) treated with 200ug/kg PO of Ivermectin per day for 2-3 days along with 100mg PO doxycycline twice per day for 5-10 days plus standard therapy; the second arm is 70 COVID-19 patients (48 mild-moderate and 22 severe and zero critical patients) on standard therapy. The time to recovery, the progression of the disease, and the mortality rate were the outcome-assessing parameters.

**Results:** among all patients and among severe patients, 3/70 (4.28%) and 1/11 (9%), respectively progressed to a more advanced stage of the disease in the Ivermectin-Doxycycline group versus 7/70 (10%) and 7/22 (31.81%), respectively in the control group (P>0.05). The mortality rate was 0/48 (0%), 0/11 (0%), and 2/11 (18.2%) in mild-moderate, severe, and critical COVID-19 patients, respectively in Ivermectin-Doxycycline group versus 0/48 (0%), and 6/22 (27.27%) in mild-moderate and severe COVID-19 patients, respectively in standard therapy group (p=0.052). Moreover, the mean time to recovery was 6.34, 20.27, and 24.13 days in mild-moderate, severe, and critical COVID-19 patients, respectively in Ivermectin-Doxycycline group versus 13.66 and 24.25 days in mild-moderate and severe COVID-19 patients, respectively in standard therapy group (P<0.01).

**Conclusions:** Ivermectin with doxycycline reduced the time to recovery and the percentage of patients who progress to more advanced stage of disease; in addition, Ivermectin with doxycycline reduced mortality rate in severe patients from 22.72% to 0%; however, 18.2% of critically ill patients died with Ivermectin and doxycycline therapy. Taken together, the earlier administered Ivermectin with doxycycline, the higher rate of successful therapy.

## Introduction

Since December 2019, the world has been facing unprecedented health caused by global spread of a novel coronavirus, SRARS-CoV-2 which causes respiratory, and multi-organ viral infection, called COVID-19. The pandemic of COVID-19 resulted in 35 million infections and more than one million deaths till the date of writing this article [1]. Almost all patients start as mild-moderate disease; however, about 15% of them progress within 5-14 days post infection to a more advanced stage of the disease, being severe and critical patients [2]. The highest risk patients have been shown to be elderly, obese, diabetic, immunosuppressed, or those with cardiovascular diseases [3, 4].

The main problem of COVID-19, there is no curative therapy till now. And the vaccine development takes time. Even after developing a successful vaccine, SARS-CoV-2 seems to induce a short lasting humeral immunity, 3-12 months only. Therefore, in the coming years when COVID-19 will be converted to seasonal endemic viral infection, a reliable therapy is still needed along with working vaccines [5].

From December 2019 till September 2020, no single drug was found to be a silver bullet for COVID-19. COVID-19 starts as respiratory viral infection but in some patients progress to severe viral pneumonia and very dangerous immune deregulation condition called cytokine storm which if is not promptly treated, it might result in acute respiratory distress syndrome (ARDS), multi-organ failure, lethal coagulopathies and death [2, 4, 6]. Therefore, potent antiviral therapy and immunomodulatory therapy for COVID-19 patients are desperately needed.

Ivermectin reduced viral load of SARS-CoV-2 *in vitro* by 5000 folds within 48h [7]. Moreover, several previous reports revealed antiviral activity of Ivermectin on Dengue HIV, Yellow fever, West Nile, Hendra, Newcastle, and Zika viruses [8-10]. Furthermore, several observational studies and real-world clinical practice showed that Ivermectin is effective in treating COVID-19 patients at both mild-moderate and severe phases of the disease [11-14]; accordingly, it is thought that Ivermectin might possess antiviral as well as immunomodulatory activity [12-14]. Ivermectin is most probably a host-specific antiviral drug and it acts as a specific inhibitor of importin α/β-mediated nuclear import inhibiting replication of several viruses such as HIV-1, Zika and dengue viruses [10]. It is thought that Ivermectin might inhibit SARS-CoV-2 using the same mechanism [7]. In addition, there have been several reports revealed that Ivermectin acts as anti-inflammatory and immunomodulatory agent and it can curb over-reacting innate and cellular immune responses [8, 15]. This explains how Ivermectin could alleviate symptoms of COVID-19 patients at viral replication phase (the first 7-10 days of infection) as well as the later hyperinflammatory phase [7-10].

Doxycycline is a broad spectrum antibiotic with reported antiviral activities on several viruses including SARS-CoV-2 [16-18]. The mechanism of the antiviral effects of tetracycline derivatives might be due to transcriptional upregulation of intracellular zinc finger antiviral protein which serves for encoding genes in host cells [19]. Doxycycline acts as an ionophore for zinc facilitating zinc entry into human cells and increasing cytoplasmic zinc concentration; high intracellular zinc levels are inhibitory to the replication of RNA viruses in cytoplasm of the cells by inhibiting RNA-depndent RNA polymerase enzyme [20]. Moreover, doxycycline has immune dampening effect making it useful to ease over-reacting immune systems [21].

In an attempt to find an effective therapy to COVID-19 patients, the current clinical trial was set up to test the combinational therapy of Ivermectin and Doxycycline in treating COVID-19 patients at different stages of the disease.

## Patients and methods

### Patients

One hundred forty (140) COVID-19 patients at different stages of the disease were included in this study. Half of them (70 patients) received Ivermectin with Doxycycline and standard care while the other half (70 patients) received the standard care only. The patients were recruited in Alkarkh and Alforat hospitals in Baghdad city in the duration from July 1^st^ to September 30^th^. The recruited patients were either outpatients or inpatients, according to the severity of the disease. Mild-moderate patients were outpatients while severe and critical patients were all inpatients. All of the recruited COVID-19 patients were diagnosed by clinical, radiological and laboratory PCR testing. Alike, recovery of COVID-19 patients was based on the disappearance of symptoms, clearance of radiological chest x-ray or Ct-scans, and getting negative PCR results.

The classification of COVID-19 patients to mild-moderate, severe, and critical was carried out according to the WHO guidelines. Ivermectin-Doxycycline group consisted of 48 mild-moderate, 11 severe and 11 critical patients while the control group consisted of 48 mild-moderate and 22 severe patients. For ethical basis, no critical patient recruited in this study was allocated to the control group; all of critical patients were allocated to the Ivermectin-Doxycycline group. The classification of the recruited patients was based on the stage of the disease at the first day of recruitment; the designated therapy of the current study was initiated at the first day of recruitment. Inclusion criteria of the patients enrolled in the clinical trial were those who were symptomatic for no more than three days for mild-moderate cases, no more than two days after being severe cases, and no more than one day after being critical cases. The purpose behind this was to assess Ivermectin-Doxycycline therapy versus standard care therapy at the beginning of each stage of the disease. The recruited patients were monitored till recovery or death.

The present study was approved by the ethical and scientific committee in Baghdad-Alkarkh General Directorate of under the approval number BKH-CT-016.

### Randomization of patients

COVID-19 patients were randomly allocated to one of the study groups depending on a simple method. Patients recruited at dates with odd number were allocated to Ivermectin-Doxycycline group while other patients were allocated to the control group. Inside each group, maximal limit of 48 mild-moderate patients and 22 severe and/or critical patients were allowed. The randomization process as well as the patients records for disease progression, recovery, and clinical or laboratory testing were supervised by the health authority of Alkarkh Health General Directorate in Baghdad city.

### Protocols of therapy

#### Ivermectin-Doxycycline group

Ivermectin 200ug/kg PO per day for two days, and in some patients who needed more time to recover, a third dose 200ug/kg PO per day was given 7 days after the first dose. Doxycycline 100mg capsule PO every 12h per day was given for 5-10 days, based on the clinical improvement of patients. In addition, standard care was given to the patients of Ivermectin-Doxycycline group based on the clinical condition of each patient.

Control group: The patients in this group received only standard care which included all or some of the following, according to the clinical condition of each patient.

#### Standard care

- Acetaminophen 500mg on need
- Vitamin C 1000mg twice/ day
- Zinc 75-125 mg/day
- Vitamin D3 5000IU/day
- Azithromycin 250mg/day for 5 days
- Oxygen therapy/ C-Pap if needed
- Dexamethazone 6 mg/day or methylprednisolone 40mg twice per day, if needed
- Mechanical ventilation, if needed

### Outcome-assessing parameters of the disease progression or recovery

Three parameters used in the present study were to assess the disease progression or recovery in COVID-19 patients who received standard care only compared to patients who received standard care with Ivermectin and Doxycycline therapy. These three parameters are:

1. Time to recovery, if any. It is the time between taking therapy till recovery.
2. Percentage of patients who progress to a more advanced stage of the disease after at least 3 days of giving therapy. For example, a patient was recruited as mild-moderate; after 3 days from starting therapy, the patient progressed to severe stage; such patient is considered a progressing patient even though she or he was under treatment.
3. Mortality rate among mild-moderate, severe, or critical patients in Ivermectin-Doxycycline group versus those in control group.

### Statistical analysis

Data was processed according to the normality tests results; parametric data were represented with mean values and non-parametric data were represented with median values. Percentage of mortality rate was calculated for each group of the study. Odds ratio and chi-square were used to test the strength and significance of association. P values less than 0.05 was considered significant

## Results

### Patients characteristics

Mean age of the recruited patients was 48.7±8.6 year with range 16 to 86 year; patients in Ivermectin-Doxycycline and control groups were age- and sex-matched. Mean age of Ivermectin-Doxycycline group was 50.1±9.3 year with 53% males and 47% females while mean age of control group patints was 47.2±7.8 year with 51% male and 49% females (P>0.05). In both groups, the median post-infection day for starting therapy was 3 days in mild-moderate, 7 days in severe, and 8.5 days in critical cases. The mean weight of Ivermectin-Doxycycline and control patients was 79.6±13.2 kg and 71.5±11.9 Kg, respectively (P>0.05).

### Time to recovery

The time to recovery was shown to be significantly reduced in the Ivermectin-Doxycycline compared to the control group; mean recovery time in Ivermectin-Doxycycline group was 10.61± 5.3 days versus mean recovery time in control group, 17.9±6.8 days (P<0.05). Hence, using Ivermectin along with Doxycycline reduced mean time to recovery up to 7 days. By analyzing the mean time to recovery in mild-moderate, severe, or critical patients in each group, it was shown that the mean time to recovery in Ivermectin-Doxycycline group was 6.34±2.4, 20.27±7.8, 19.77±9.2 days, respectively versus 13.66±6.4, 24.25±9.5 days, in control group, respectively (P<0.01). Accordingly, Ivermectin-Doxycycline reduced recovery time about 7.32 days in mild-moderate, and 3.98 or roughly 4 days in severe patients (Table 1).

**Table 1:** Parameters of the study outcomes between Ivermectin-Doxycycline and control groups

| Outcome parameter | Subgroup of patients | Ivermectin-Doxycycline | Control | P value |
| --- | --- | --- | --- | --- |
| Time to recovery (day) | Total | 10.61± 5.3 | 17.9±6.8 | <0.0001 |
|  | Mild-moderate | 6.34±2.4 | 13.66±6.4 | <0.0001 |
|  | severe | 20.27±7.8 | 24.25±9.5 | 0.29 |
|  | critical | 19.77±9.2 | / |  |
| Rate of progression of disease number/total (%) | Total | 3/70 (4.28%) | 7/70 (10%) | 0.19 (OR*=0.4, P=0.2) |
|  | Mild-moderate | 0/48 (0%) | 0/48 (0%) | 1 |
|  | severe | 1/11 (9%) | 7/22 (31.81%) | 0.15 (OR=0.21, P=0.17) |
|  | critical | 2/11 (18.2%) | / |  |
| Mortality rate | Total | 2/70 (2.85%) | 6/70 (7.14) | 0.14 (OR=0.31, P=0.16) |
|  | Mild-moderate | 0/48 (0%) | 0/48 (0%) | 1 |
|  | severe | 0/11 (0%) | 6/22 (27.27%) | 0.052 (OR=0.11, P=0.14) |
|  | critical | 2/11 (18.2%) | / |  |

OR\*: Odds ratio

### Progression of the disease

The rate of progression of the disease, or the deterioration of the clinical condition of the patients despite of taking standard care with/without Ivermectin-doxycycline therapy, was shown to be varied between the two groups studied. At beginning, no single mild-moderate patient in both groups progressed to a more advanced stage of the disease. For severe COVID-19 patients, 1/11 (9%) in Ivermectin-Doxycycline group versus 7/22 (31.81%) in control group progressed to more advanced stage of the disease, namely being classified as critical cases (P>0.05). Thus, Ivermectin-Doxycycline protocol was shown to lower progression of the disease in severe patients if given within the first two days of the severe stage of the disease. For critical patients, ethically, critical patients were not included in the control group as critical patients need to receive all possible medications for saving their lives; hence, it was not possible to compare the rate of progression of the disease in critical patients between the two studied groups (Table 1).

### Mortality rate

The mortality rate was shown to be 0/48 (0%) in mild-moderate patients in both groups. Nevertheless, the mortality rate was diminished to 0/11 (0%) in Ivermectin-Doxycycline group compared to 6/22 (27.27%) (P=0.052). For critical patients, Ivermectin-Doxycycline did not prevent death in those patients as mortality rate was shown to be 2/11 (18.2%), (Table 1). No critical patients were included in the control group to compare with; however, it is obvious that 18.2% of death in critical COVID-19 patients received Ivermectin-Doxycycline is much lower than the death rate in critical cases of COVID-19 in Iraq that might reach above 50% (based on real-world data not official published data).

## Discussion

Finding an effective therapy for COVID-19 is an ultimate goal for health bodies all over the world. The problem of the standard care for COVID-19 patient is not curative; however, the current situation is much better than the first months of the pandemic, after introducing steroid therapy for severe/critical patients and high doses of vitamin D3, vitamin C and Zinc for mild-moderate cases [22]. COVID-19 is a multiphasic disease starting with virus replicative phase lasting for 7-10 days then, in some patents, is followed with hyperinflammatory phase and cytokine storm where the most fatalities occur [2, 4]. If the viral and hyperinflammatory phases of the disease were not addressed early and properly, the patient might progress to ARDS which is almost fatal [2,6]. Hence, antiviral, anti-inflammatory, and immunomodulatory medications are necessary to stop the vicious progression of COVID-19 from mild-moderate to severe and to stop the clinical deterioration of the already severe patients.

Accordingly, Ivermectin and Doxycycline were used in this study because both drugs have shown antiviral and immunomodulatory activities [8-15]. Moreover, Doxycycline is a broad spectrum antibiotic which tackles the problem of secondary bacterial infection in COVID-19 patients [16-20]. Both drugs are FDA approved and have a high historical safety record [8,10,16, 19]; moreover, no interaction is known between Ivermectin and Doxycycline or between Ivermectin-Doxycycline and any of the medications given in the standard care. By contrary, for example, Azithromycin with Hydroxychloroquine are known to interact adversely for prolonging QT interval of cardiogram which might lead to serious complications [23].

Using FDA approved and safe antiviral and immunomodulatory medications for COVID-19 is scientifically justified for untreatable disease like COVID-19. It has been found that most of COVID-19 patients who progress to severe/critical disease have high viral load of SARS-CoV-2 and over-reacting immune response [24]. Therefore, reducing the viral load and dampening the immune response and inflammatory cytokines are necessary to save patients’ lives.

The findings of the current trial showed that Ivermectin-Doxycycline reduced the mean time to recovery from 17.9 to 10.61 days in the recruited COVID-19 patients. Alike, for mild moderate patients, Ivermectin-Doxycycline reduced mean time to recovery from 13.66 to just 6.34 days with reduction in time up to 7.32 days. Nevertheless, Ivermectin-Doxycycline reduced the mean time to recovery in severe patients only 4 days, from 24 to 20 days. Based on these findings, Ivermectin and Doxycycline protocol proves to be effective in speeding up recovery in both mild-moderate outpatients and severe inpatients. This has a tremendous effect on lowering the burden of the disease, minimizing chances of developing immune deregulation, and freeing as quickly as possible hospital beds to other patients. This adds further evidence that Ivermectin-Doxycycline could exert both antiviral and immunomodulatory actives. Several observational studies showed that Ivermectin with/without Doxycycline shortens the time needed to recover COVID-19 patients and Ivermectin is beneficial for mild-moderate as well as severe patients [8-15].

In the current study, Ivermectin-Doxycycline arm lowered the rate of progression of the severe patients from 31.81% to as low as 9%. More interestingly Ivermectin-Doxycycline abolished death in severe patients, 0% mortality rate, compared to control arm, 27.27%. It is noteworthy to mention that the non-progression of the disease and the zero mortality in mild-moderate patients in both arms of the study might be attributed to the early diagnosis and therapy; moreover, the current standard care has become more effective than that used in the early months of the pandemic. However, larger study population is required to trace differences in the disease progression or the mortality rate of mild-moderate patients of COVID-19 taking Ivermectin-Doxycycline compared to patients taking standard care.

Accordingly, the present clinical trial reveals that Ivermectin-Doxycycline might stop disease progression and reduce death rate in severe patients of COVID-19. An observational preprint study conducted in Florida showed that Ivermectin cuts mortality rate of severe COVID-19 patients from 80.7% to 38.8% [25]. Interestingly, both Ivermectin and Doxycycline concentrations in the tissue of the lung have been estimated 2 times more than that in plasma [26, 27]. Therefore, their antiviral and anti-inflammatory effect on pulmonary tissues is expected to be prominent. These findings provide evidence that Ivermectin might be a potent immunomodulatory in addition to being antiviral agent. Nevertheless, these observational findings still need confirmation by a large randomized controlled study.

In fact, the observed benefits of Ivermectin-Doxycycline on the enrolled COVID-19 patients cannot be separated from the effect of the standard care, including Vitamin D and C, Zinc, and steroids, which was given concomitantly. Therefore, giving Ivermectin-Doxycycline along with Zinc, Vitamins D and C, and steroids at the viral and/or the hyperinflammatory phase of the disease seems clinically of benefit. This might shape the future most-fit combinational therapy of COVID-19 patients to minimize as could as possible the death rate and to decrease the duration and the progression of the disease.

## Data Availability

All data are available

## References

1. Worldmeter. Coronavirus update, October 6th. https://www.worldometers.info/coronavirus/?

2. Kuldeep Dhama, Sharun Khan, Ruchi Tiwari, Shubhankar Sircar, Sudipta Bhat, Yashpal Singh Malik, Karam Pal Singh, Wanpen Chaicumpa, D. Katterine Bonilla-Aldana, Alfonso J. Rodriguez-Morales. Coronavirus Disease 2019–COVID-19. Clinical Microbiology Reviews Jun 2020, 33 (4) e00028–20; DOI: 10.1128/CMR.00028-20.

3. Rod J.E., Oviedo-Trespalacios Oscar, Cortes-Ramirez Javier. A brief-review of the risk factors for covid-19 severity. Rev. Saúde Pública [Internet]. 2020 [cited 2020 Oct 08] ; 54: 60. Available from: http://www.scielo.br/scielo.php?script=sci_arttext&pid=S0034-89102020000100701&lng=en. Epub June 01, 2020. http://dx.doi.org/10.11606/s1518-8787.2020054002481.

4. Abdulamir AS, Hafidh RR. The Possible Immunological Pathways for the Variable Immunopathogenesis of COVID—19 Infections among Healthy Adults, Elderly and Children. Electron J Gen Med. 2020;17(4):em202. https://doi.org/10.29333/ejgm/7850

5. Huang AT, Garcia-Carreras B, Hitchings MDT, et al. A systematic review of antibody mediated immunity to coronaviruses: antibody kinetics, correlates of protection, and association of antibody responses with severity of disease. Preprint. medRxiv. 2020;2020.04.14.20065771. Published 2020 Apr 17. doi:10.1101/2020.04.14.20065771

6. Rasheed AM, Fatak DF, Hashim HA, Maulood MF, Kabah KK, Almusawi YA, Abdulamir AS. The therapeutic potential of convalescent plasma therapy on treating critically-ill COVID-19 patients residing in respiratory care units in hospitals in Baghdad, Iraq. Infez Med. 2020 Sep 1;28(3):357–366. PMID: 32920571.

7. Leon Caly, Julian D. Druce, Mike G. Catton, David A. Jans, Kylie M. Wagstaff. The FDA-approved drug ivermectin inhibits the replication of SARS-CoV-2 in vitro. Antiviral Research. Volume 178, June 2020, 104787.

8. Heidary, F., Gharebaghi, R. Ivermectin: a systematic review from antiviral effects to COVID-19 complementary regimen. J Antibiot 73, 593–602 (2020). https://doi.org/10.1038/s41429-020-0336-z

9. Xu TL, Han Y, Liu W, et al. Antivirus effectiveness of ivermectin on dengue virus type 2 in Aedes albopictus. PLoS Negl Trop Dis. 2018;12(11):e0006934. Published 2018 Nov 19. doi:10.1371/journal.pntd.0006934.

10. Wagstaff KM, Sivakumaran H, Heaton SM, Harrich D, Jans DA. Ivermectin is a specific inhibitor of importin α/β-mediated nuclear import able to inhibit replication of HIV-1 and dengue virus. Biochem J. 2012;443(3):851–856. doi:10.1042/BJ20120150.

11. Vora A, Arora VK, Behera D, Tripathy SK. White paper on Ivermectin as a potential therapy for COVID-19. Indian J Tuberc. 2020;67(3):448–451. doi:10.1016/j.ijtb.2020.07.031.

12. Mudatsir, M.; Yufika, A.; Nainu, F.; Frediansyah, A.; Megawati, D.; Pranata, A.; Mahdani, W.; Ichsan, I.; Dhama, K.; Harapan, H. Antiviral Activity of Ivermectin Against SARS-CoV-2: An Old-Fashioned Dog with a New Trick—A Literature Review. Sci. Pharm. 2020, 88, 36.

13. Hector Eduardo Carvallo, Roberto Raul Hirsch, Maria Eugenia Farinella. Safety and Efficacy of the combined use of ivermectin, dexamethasone, enoxaparin and aspirin against COVID-19. medRxiv 2020.09.10.20191619 ; Preprint doi: https://doi.org/10.1101/2020.09.10.20191619

14. Faiq I. Gorial, Sabeeh Mashhadani, Hend M Sayaly, Basim Dhawi Dakhil, Marwan M AlMashhadani, Adnan M Aljabory,, Hassan M Abbas, Mohammed Ghanim, Jawad I Rasheed. Effectiveness of Ivermectin as add-on Therapy in COVID-19 Management. medRxiv 2020.07.07.20145979; Preprint doi: https://doi.org/10.1101/2020.07.07.20145979

15. Zheng, H. J., Tao, Z. H., Cheng, W. F., Wang, S. H., Cheng, S. H., Ye, Y. M., Luo, L. F., Chen, X. R., Gan, G. B. and Piessens, W. F. (1991). Efficacy of ivermectin for the control of microfilaremia recurring after treatment with diethylcarbamazine. II. Immunological changes following treatment. American Journal of Tropical Medicine and Hygiene 45, 175–181.

16. Alexandre E.MalekBruno P.GranwehrDimitrios P. Kontoyiannis. Doxycycline as a potential partner of COVID-19 therapies. IDCases-Volume 21, 2020, e00864.

17. Mathieu Gendrota, Julien Andreanic, Priscilla Jardotc, Sébastien Hutter, Manon Boxberger, Joel Mosnier, Marion Le Bideau, Isabelle Duflot, Isabelle Fonta, Clara Rolland, Hervé Bogreau, Bernard La Scola, Bruno Pradines. In vitro antiviral activity of doxycycline against SARS-CoV-2. Preprint: Mediteranee Infection website: https://www.mediterranee-infection.com/in-vitro-antiviral-activity-of-doxycycline-against-sars-cov-2/

18. Wu ZC, Wang X, Wei JC, Li BB, Shao DH, Li YM, Liu K, Shi YY, Zhou B, Qiu YF, Ma ZY. Antiviral activity of doxycycline against vesicular stomatitis virus in vitro. FEMS Microbiol Lett. 2015 Nov;362(22):fnv195. doi: 10.1093/femsle/fnv195. Epub 2015 Oct 12. PMID: 26459887.

19. M.J. Bick, J.-W.N. Carroll, G. Gao, S.P. Goff, C.M. Rice, M.R. MacDonald. Expression of the zinc-finger antiviral protein inhibits alphavirus replication J Virol (2003), 10.1128/jvi.77.21.11555-11562.2003.

20. Ali I, Alfarouk KO, Reshkin SJ, Ibrahim ME. Doxycycline as Potential Anti-cancer Agent. Anticancer Agents Med Chem. 2017;17(12):1617–1623. doi: 10.2174/1871520617666170213111951. PMID: 28270076.

21. J.Z. Castro, T. Fredeking. Doxycycline modify the cytokine storm in patients with dengue and dengue hemorrhagic fever. Int J Infect Dis (2010), 10.1016/j.ijid.2010.02.1586.

22. Ali MJ, Hanif M, Haider MA, et al. Treatment Options for COVID-19: A Review. Front Med (Lausanne). 2020;7:480. Published 2020 Jul 31. doi:10.3389/fmed.2020.00480.

23. Thomas F. O’Connell, Christopher J. Bradley, Amr E. Abbas, Brian D. Williamson, Akash Rusia, Adam M. Tawney, Rick Gaines, Jason Schott, Alex Dmitrienko, David E. Haines. Hydroxychloroquine/Azithromycin Therapy and QT Prolongation in Hospitalized Patients with COVID-19. Am Coll Cardiol EP. 2020 Aug 05. Epublished DOI:10.1016/j.jacep.2020.07.016.

24. Pujadas E, Chaudhry F, McBride R, Richter F, Zhao S, Wajnberg A, Nadkarni G, Glicksberg BS, Houldsworth J, Cordon-Cardo C. SARS-CoV-2 viral load predicts COVID-19 mortality. Lancet Respir Med. 2020 Sep;8(9):e70. doi: 10.1016/S2213-2600(20)30354-4. Epub 2020 Aug 6. PMID: 32771081.

25. Juliana Cepelowicz Rajter, Michael Sherman, Naaz Fatteh, Fabio Vogel, Jamie Sacks, Jean-Jacques Rajter. ICON (Ivermectin in COvid Nineteen) study: Use of Ivermectin is Associated with Lower Mortality in Hospitalized Patients with COVID19. MedrxIV preprint: doi: https://doi.org/10.1101/2020.06.06.20124461.

26. Vargas-Estrada D, Gutirrez L, Juarez-Rodriguez I, Sumano H. Pharmacokinetics of doxycycline and tissue concentrations of an experimental long-acting parenteral formulation of doxycycline in Wistar rats. Arzneimittelforschung 2008; 58: 310–5.

27. Lifschitz A, Virkel G, Sallovitz J, Sutra JF, Galtier P, Alvinerie M, Lanusse C. Comparative distribution of ivermectin and doramectin to parasite location tissues in cattle. Vet Parasitol. 2000 Feb 1;87(4):327–38. doi: 10.1016/s0304-4017(99)00175-2. PMID: 10669102

